# Prevalence of Isolated Systolic Hypertension among People with Diabetes in Indonesia

**DOI:** 10.1101/2020.11.25.20238683

**Authors:** Mahalul Azam, Fetty Nur Hidayati, Arulita Ika Fibriana, Udin Bahrudin, Syed Mohamed Aljunid

## Abstract

The present study aimed to explore the prevalence of isolated systolic hypertension (ISH) and its risk factors among diabetes mellitus (DM) subjects in the community setting study in Indonesia. This cross-sectional study extracted secondary data from basic health survey (Riset Kesehatan Dasar; RISKESDAS) conducted in 2018. DM subjects were defined based on fasting blood glucose level ≥ 126 mg/dL or 2 hours postprandial and random blood glucose level ≥ 200 mg/dL or previously had been diagnosed by a doctor; while ISH was determined based on systolic blood pressure ≥ 140 mmHg and diastolic blood pressure < 90 mmHg. We also observed the subject’s characteristics, such as demography, lipid profile, and subject’s compliance. Data were then analyzed using Chi-square and Binary logistic regression. Study involved 3,911 DM subjects, revealed the overall prevalence of ISH 17.5%. Age category of 35-44 years old (POR= 10.80; 95%CI: 2.595-44.957), 45-54 years old (POR=22.81; 95%CI: 5.616-92.677), 55-64 years old (POR=46.12; 95% CI: 11.393-186.720); 65-74 years old (POR= 81.82; 95% CI: 20.110-332.868); ≥75 years old (POR=109.64; 95% CI: 26.373-455.789), low HDL cholesterol (POR= 0,80; 95% CI: 0.653-0.972); duration of DM (POR= 1.73; 95% CI: 1.257-2.389) were associated with the ISH. The prevalence of ISH among DM subjects was 17.5%. Older DM subjects, low HDL cholesterol, and duration of DM were associated with the ISH, suggesting that modification lipid profile, especially the HDL cholesterol level, is an important measure to delay ISH in elderly and long-duration DM subjects.

## Introduction

International Diabetes Federation reports 463 million people globally, and 10.7 million people in Indonesia living with diabetes placing Indonesia in the 7^th^ rank among countries for the number of adults with diabetes [1]. Hypertension is the most frequent comorbidity for diabetes [2–4]. Both hypertension and diabetes are the major risk factors for cardiovascular diseases due to the vascular mechanism [5]. Hypertension is associated with 30% of death and 25% of cardiovascular events among diabetes mellitus (DM) subjects [6]. DM subjects with hypertension have seven times likely to experience end-stage renal disease and 2-4 times to get myocardial infarction and stroke [6].

Hypertension occurred due to the vascular resistance and increase of fluid volume [7]. Vascular resistance in DM subjects is related to vascular remodeling that caused arterial stiffness, while the increase of body fluid volume is associated with resistance-induced hyperinsulinemia and hyperglycemia [7]. Isolated systolic hypertension (ISH) is the most frequent form of hypertension among the elderly [8] and the most frequent subtype of uncontrolled hypertension [9]. People with diabetes have twice higher risk to get ISH than of those without diabetes [10]. ISH reflects widespread atherosclerosis and increases stroke risk of 11% as well as an increase in all-cause mortality risk of 16% [10]. Alongside the ISH, the pulse pressure (PP) and mean arterial pressure (MAP) is the independent predictors of cardiovascular events and all-cause mortality [10–13].

A previous study[14] based on the hospital-based data reported that the prevalence of ISH among DM subjects was 37.4%, and age was the most related factor. Another study reported that the prevalence of ISH among DM subjects was 27.6%[15]; male, older age, obesity, and smoking were its risk factors [15,16]. A study in Indonesia reported risk factors of hypertension among DM subjects such as age, mental health disorders, obesity, physical activities, duration of diabetes, dyslipidemia, and patient compliance [17]. However, limited information regarding prevalence and risk factors of ISH among DM subjects based on population-based data. The present study aimed to explore the prevalence of ISH and its risk factors among DM subjects based on community setting study in Indonesia.

## Materials and Methods

### Design and study population

This cross-sectional study extracted secondary data from the basic health survey (Riset Kesehatan Dasar; RISKESDAS) 2018, the latest five-annual national scope cross-sectional study, conducted by the National Institute of Research and Development, Ministry of Health, the Republic of Indonesia. The survey was conducted and delivered for households systematic-randomly selected from 514 districts/cities in 34 provinces. For each province and district/city, the number of proportional census blocks was determined systematically. Three hundred households or 30.000 census blocks were then determined to be involved in the survey. Of them, 94.2 % or 282,654 households completed the questionnaire consist of 1,017,290 individual subjects[18]. The study population involved subjects with DM in the RISKESDAS 2018 data. Subjects with DM were determined by fasting blood glucose level ≥ 126 mg/dL or 2 hours postprandial and random blood glucose level ≥ 200 mg/dL or previously had been diagnosed by a doctor.

### Data collection

Ethical clearance for the RISKESDAS 2018 study was obtained from the Ethics Committee, the National Institute of Health Research and Development (NIHRD), the Ministry of Health, Republic of Indonesia. Subject with ISH was defined as those with systolic blood pressure ≥ 140 mmHg and diastolic blood pressure < 90 mmHg [19]. We categorized the subject as non-hypertensive when meet the criteria of optimal (<120 mmHg and <80 mmHg), or normal (120 mmHg-129 mmHg and/or 80-84 mmHg), or high normal (130-139 mmHg and/or 85-89 mmHg). While non ISH hypertension were categorized for grade 1-3 hypertension; grade 1 hypertension: 140-159 mmHg and/or 90-99 mmHg; grade 2 hypertension: 160-179 mmHg and/or 100-109 mmHg; grade 3 hypertension: ≥180 mmHg and or ≥110 mmHg [19]. Based on the measurement of blood pressure, we also calculated pulse pressure (PP) and mean arterial pressure (MAP). PP was calculated as a result of the formula (PP = systolic blood pressure (SBP) – diastolic blood pressure (DBP)), while the MAP was calculated as the formula of 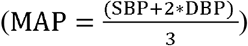.

Secondary data acquired from RISKESDAS 2018 were age, sex, urban-rural residence status, marital status, educational level, employment status, total cholesterol level, HDL-cholesterol level, triglycerides level, history of hypertension, smoking, physical activity status, alcohol consumption, body mass index (BMI), duration of DM, type of medication, and medication compliance.

### Statistical analysis

Characteristics of the subjects were presented as proportions since they are categorical type of data. The association between ISH status were analyzed using the Chi-square test. The p-values <0.05 were considered statistically significant. Parameters that had p-value <0.25 were then involved in the multivariate analysis using binary logistic regression. All statistical analyses were performed using the Statistical Package for the Social Sciences (SPSS) software (version 23.0 for Windows, IBM SPSS Inc., Chicago, IL).

## Results

Data extracted from the RISKESDAS 2018 consisted of 3,911 DM subjects that were included in the final analysis. Study population consisted of 1,289 (33%) male and 2,622 (67%) female. The most frequent age category was 45-54 years old (29.3 %). More than half of the study population was live in the urban area with a low level of education and were employed in various sectors. Most of the study population had lower total cholesterol levels, lower high-density lipoprotein (HDL) cholesterol level, higher low-density lipoprotein (LDL) cholesterol level, and lower triglyceride levels. Most of them had a history of hypertension, non-smoking, active physical activity, and fair medication compliance. The detailed subjects’ characteristics are presented in Table 1.

**Table 1.** Subjects’ characteristics

| Variable | n | % |
| --- | --- | --- |
| **Age category (years old)** |  |  |
| ≥ 75 | 167 | 4.3 |
| 65-74 | 502 | 12.8 |
| 55-64 | 1098 | 28.1 |
| 45-54 | 1146 | 29.3 |
| 35-44 | 668 | 17.1 |
| 15-34 | 330 | 8.4 |
| **Sex** |  |  |
| Female | 2622 | 67 |
| Male | 1289 | 33 |
| **Residence status** |  |  |
| Urban | 2057 | 52.6 |
| Rural | 1854 | 47.4 |
| **Marital status** |  |  |
| Un-married | 147 | 3.8 |
| Married | 3764 | 96.2 |
| **Educational level** |  |  |
| Primary high school | 3006 | 76.9 |
| Secondary high school or above | 905 | 23.1 |
| **Employment status** |  |  |
| Un-employed | 1639 | 41.9 |
| Employed | 2272 | 58.1 |
| **Total Cholesterol level** |  |  |
| ≥200 mg/dL | 1832 | 46.8 |
| <200 mg/dL | 2079 | 53.2 |
| **HDL level** |  |  |
| ≥40 mg/dL | 1140 | 29.1 |
| <40 mg/dL | 2771 | 70.9 |
| **LDL level** |  |  |
| ≥100 mg/dL | 3296 | 84.3 |
| <100 mg/dL | 615 | 15.7 |
| **Triglyceride** |  |  |
| ≥150 mg/dL | 1612 | 41.2 |
| <150 mg/dL | 2299 | 58.8 |
| **History of hypertension** |  |  |
| Yes | 1350 | 34.5 |
| No | 2561 | 65.5 |
| **Smoking** |  |  |
| Yes | 1002 | 25.6 |
| No | 2909 | 74.4 |
| **Physical activity status** |  |  |
| Sedentary | 605 | 15.5 |
| Active | 3306 | 84.5 |
| **Alcohol consumption** |  |  |
| Yes | 37 | 0.9 |
| No | 3874 | 99.1 |
| **BMI** |  |  |
| Obese | 1262 | 32.3 |
| Overweight | 603 | 15.4 |
| Normal | 1767 | 45.2 |
| Underweight | 164 | 4.2 |
| Severe Underweight | 115 | 2.9 |
| **Duration of DM** |  |  |
| >5 years | 1867 | 47.7 |
| <5 years | 2044 | 52.3 |
| **Type of DM medication** |  |  |
| No medication | 830 | 21.2 |
| OHD+insulin | 828 | 21.2 |
| Insulin | 770 | 19.7 |
| OHD | 1483 | 37.9 |
| **Medication compliance** |  |  |
| No | 1625 | 41.5 |
| Yes | 2286 | 58.5 |

BMI: body mass index; HDL=high-density lipoprotein; LDL=low-density lipoprotein

Based on the result of blood pressure measurement, a total of 1,903 (48.7%) subjects were categorized as normal, while the rest of 2,008 (51.3%) were categorized as hypertension, whether grade 1,2,3 or ISH. ISH was the most frequent form of hypertension in the study population (Fig. 1). The highest mean MAP was in non-ISH hypertension group, while the highest mean PP was in the ISH group (Fig. 2)

**Figure 1.**
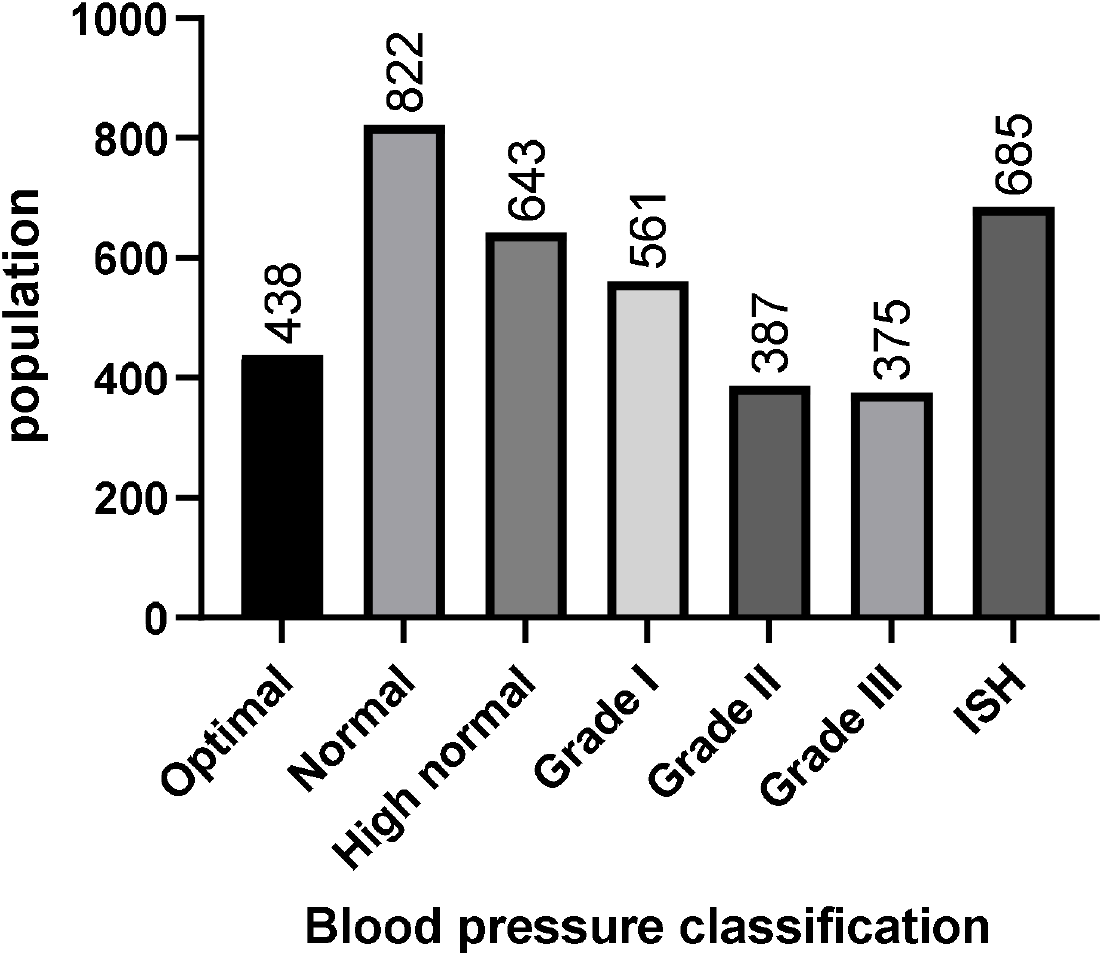
Frequency of blood pressure classification among DM subjects Optimal: <120 and <80; Normal: 120-129 and/or 80-84; High normal: 130-139 and/or 85-89; Grade I hypertension: 140-159 and/or 90-99; Grade II hypertension: 160-179 and or 100-109; Grade III hypertension: ≥180 and or ≥110; Isolated systolic hypertension (ISH): ≥140 and <90

**Figure 2.**
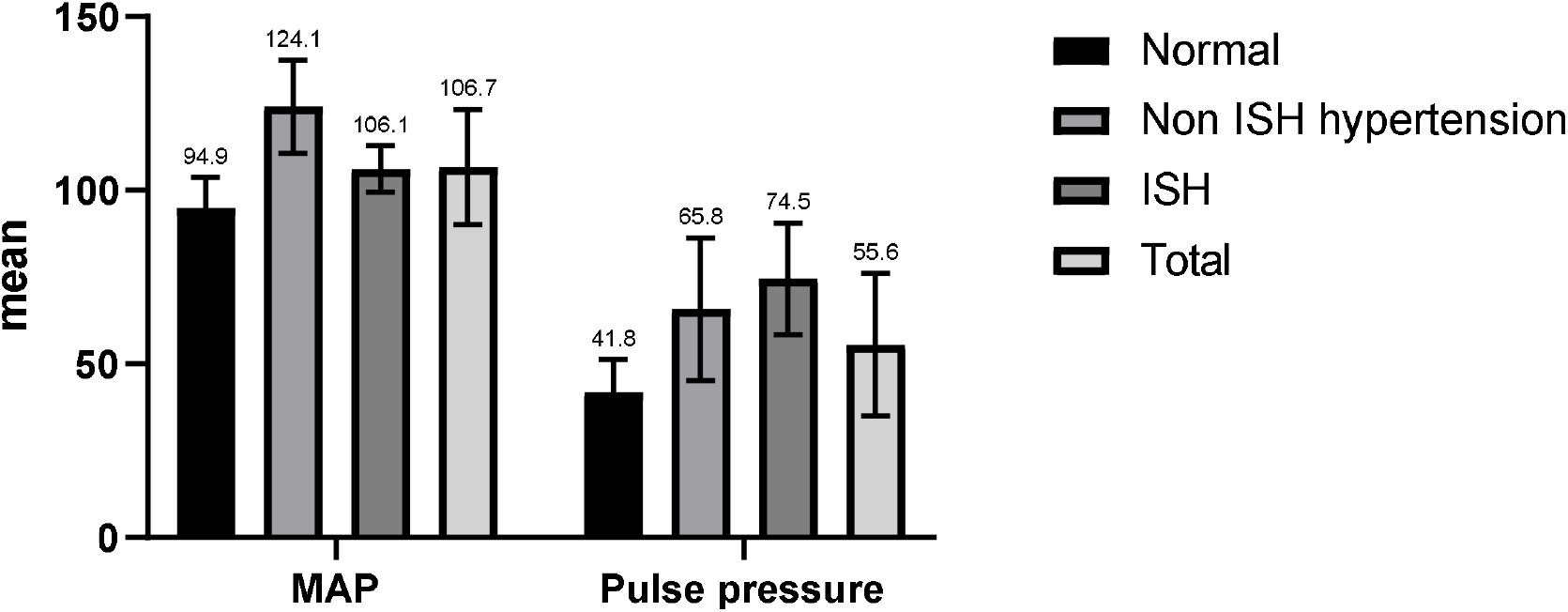
Mean arterial pressure (MAP) and pulse pressure based on hypertension classification. ISH: isolated systolic hypertension

Of the total 3,911 study population of DM subjects, 685 subjects were identified as ISH, indicated that the prevalence of ISH was 17.5%. Table 2 identified variables associated with the ISH. Older subjects, low educational level, high total cholesterol level, low HDL level, active physical activity, obese, duration of DM, and type of medication were associated with the ISH status among DM subjects. These variables, combined with other variables that p ≤ 0.25, i.e., employment status, LDL level, triglyceride, history of hypertension, alcohol consumption, and medication compliance, continued to be involved in the Binary logistic regression, and the final model of regression showed in Table 3.

**Table 2.** Subjects’ characteristics based on ISH status

| Parameter | ISH |  |  | p<sup>*</sup> | POR | 95%CI |  |
| --- | --- | --- | --- | --- | --- | --- | --- |
|  | Yes<br>n(%) | No<br>n(%) | Total<br>n(%) |  |  | Lower | Uper |
| **Age (years old)** |  |  |  |  |  |  |  |
| ≥75 | 70 (41.9) | 97 (58.1) | 167 | 0.001 | 69.16 | 17.172 | 278.560 |
| 65-74 | 174 (34.7) | 328 (65.3) | 502 | 0.001 | 57.19 | 14.289 | 228.905 |
| 55-64 | 253 (23) | 845 (77) | 1098 | 0.001 | 38.02 | 9.508 | 152.020 |
| 45-54 | 145 (12.7) | 1001 (87.3) | 1146 | 0.001 | 20.88 | 5.200 | 83.822 |
| 35-44 | 41 (6.1) | 627 (93.9) | 668 | 0.001 | 10.13 | 2.465 | 41.612 |
| 15-34 | 2 (0.6) | 328 (99.4) | 330 | Reference | 1 |  |  |
| **Sex** |  |  |  |  |  |  |  |
| Female | 451 (17.2) | 2171 (82.8) | 2622 | 0.489 | 0.95 | 0.821 | 1.093 |
| Male | 234 (18.2) | 1055 (81.8) | 1289 |  |  |  |  |
| **Residence status** |  |  |  |  |  |  |  |
| Urban | 370 (18) | 1687 (82) | 2057 | 0.437 | 1.06 | 0.924 | 1.214 |
| Rural | 315 (17) | 1539 (83) | 1854 |  |  |  |  |
| **Marital status** |  |  |  |  |  |  |  |
| Un-married | 23 (15.6) | 124 (84.4) | 147 | 0.619 | 0.89 | 0.607 | 1.303 |
| Married | 662 (17.6) | 3102 (82.4) | 3764 |  |  |  |  |
| **Education level** |  |  |  |  |  |  |  |
| Low | 553 (18.4) | 2453 (81.6) | 3006 | 0.009 | 1.26 | 1.059 | 1.502 |
| High | 132 (14.6) | 773 (85.4) | 905 |  |  |  |  |
| **Employment status** |  |  |  |  |  |  |  |
| Un-employed | 308 (18.8) | 1331 (81.2) | 1639 | 0.081 | 1.13 | 0.988 | 1.298 |
| Employed | 377 (16.6) | 1895 (83.4) | 2272 |  |  |  |  |
| **Total Cholesterol level** |  |  |  |  |  |  |  |
| ≥200 mg/dL | 345 (18.8) | 1487 (81.2) | 1832 | 0.046 | 1.15 | 1.005 | 1.319 |
| <200 mg/dL | 340 (16.4) | 1739 (83.6) | 2079 |  |  |  |  |
| **HDL level** |  |  |  |  |  |  |  |
| ≥40 mg/dL | 169 (14.8) | 971 (85.2) | 1140 | 0.005 | 0.80 | 0.679 | 0.934 |
| <40 mg/dL | 516 (18.6) | 2255 (81.4) | 2771 |  |  |  |  |
| **LDL level** |  |  |  |  |  |  |  |
| ≥100 mg/dL | 591 (17.9) | 2705 (2.1) | 3296 | 0.127 | 1.17 | 0.961 | 1.433 |
| <100 mg/dL | 94 (15.3) | 521 (84.7) | 615 |  |  |  |  |
| **Triglyceride** |  |  |  |  |  |  |  |
| ≥150 mg/dL | 261 (16.2) | 1351 (83.8) | 1612 | 0.075 | 0.88 | 0.763 | 1.010 |
| <150 mg/dL | 424 (18.4) | 1875 (81.6) | 2299 |  |  |  |  |
| **History of hypertension** |  |  |  |  |  |  |  |
| Yes | 250 (18.5) | 1100 (81.5) | 1350 | 0.248 | 1.09 | 0.947 | 1.255 |
| No | 435 (17) | 2126 (83) | 2561 |  |  |  |  |
| **Smoking** |  |  |  |  |  |  |  |
| Yes | 182 (18.2) | 820 (81.8) | 1002 | 0.563 | 1.05 | 0.901 | 1.225 |
| No | 503 (17.3) | 2406 (82.7) | 2909 |  |  |  |  |
| **Physical activity status** |  |  |  |  |  |  |  |
| Sedentary | 131 (21.7) | 474 (78.3) | 605 | 0.004 | 1.29 | 1.091 | 1.531 |
| Active | 554 (16.8) | 2752 (83.2) | 3306 |  |  |  |  |
| **Alcohol consumption** |  |  |  |  |  |  |  |
| Yes | 2 (5.4) | 35 (94.6) | 37 | 0.084 | 0.31 | 0.080 | 1.182 |
| No | 683 (17.6) | 3191 (82.4) | 3874 |  |  |  |  |
| **BMI** |  |  |  |  |  |  |  |
| Obese | 165 (13.1) | 1097 (86.9) | 1262 | 0.003 | 0.557 | 0.389 | 0.798 |
| Overweight | 91 (15.1) | 512 (84.9) | 603 | 0.037 | 0.643 | 0.439 | 0.940 |
| Normal | 368 (20.8) | 1399 (79.2) | 1767 | 0.576 | 0.887 | 0.630 | 1.249 |
| Underweight | 34 (20.7) | 130 (79.3) | 164 | 0.690 | 0.883 | 0.566 | 1.379 |
| Severe underweight | 27 (23.5) | 88 (76.5) | 115 | Reference |  |  |  |
| **Duration of DM** |  |  |  |  |  |  |  |
| >5 years | 469 (25.1) | 1398 (74.9) | 1867 | 0.001 | 2.38 | 2.049 | 2.758 |
| <5 years | 216 (10.6) | 1828 (89.4) | 2044 |  |  |  |  |
| **Type of DM medication** |  |  |  |  |  |  |  |
| No medication | 235 (28.3) | 595 (71.7) | 830 | 0.001 | 2.09 | 1.766 | 2.471 |
| OHD+insulin | 158 (19.1) | 670 (80.9) | 828 | 0.001 | 1.41 | 1.164 | 1.703 |
| Insulin | 91 (11.8) | 679 (88.2) | 770 | 0.261 | 0.87 | 0.692 | 1.099 |
| OHD | 201 (13.6) | 1282 (86.4) | 1483 | Reference | 1 |  |  |
| **Medication compliance** |  |  |  |  |  |  |  |
| No | 307 (18.9) | 1318 (81.1) | 1625 | 0.062 | 1.14 | 0.997 | 1.310 |
| Yes | 378 (16.5) | 1908 (83.5) | 2286 |  |  |  |  |

\*Chi-square test
HDL: high density lipoprotein; LDL: low density lipoprotein; OHD: oral hypoglycaemic drugs; POR: prevalence odds ratio

**Table 3.** Binary logistic regression of ISH risk factors among DM subjects

| Variables | p | POR | 95% CI. |  |
| --- | --- | --- | --- | --- |
| Age (years old) |  |  |  |  |
| $\geq 75$ | **0.001** | 109.64 | 26.373 | 455.789 |
| 65-74 | 0.001 | 81.82 | 20.110 | 332.868 |
| 55-64 | 0.001 | 46.12 | 11.393 | 186.720 |
| 45-54 | 0.001 | 22.81 | 5.616 | 92.677 |
| 35-44 | 0.001 | 10.80 | 2.595 | 44.957 |
| High HDL cholesterol | **0.025** | 0.80 | 0.653 | 0.972 |
| History of hypertension | 0.070 | 1.183 | 0.986 | 1.418 |
| BMI |  |  |  |  |
| Obese | 0.331 | 0.769 | 0.453 | 1.306 |
| Overweight | 0.493 | 0.825 | 0.475 | 1.431 |
| Normal | 0.369 | 1.241 | 0.775 | 1.987 |
| Underweight | 0.857 | 0.946 | 0.520 | 1.722 |
| Duration of DM | **0.001** | 1.73 | 1.257 | 2.389 |

BMI: body mass index; HDL: high density lipoprotein; POR: prevalence odds ratio

We found that older subjects, low HDL cholesterol (prevalence odds ratio; POR=0.80; 95% CI: 0.653-0.972), and duration of DM (POR=1.73; 95% CI: 1.257-2.389), all together were associated with the ISH. Subjects with the older age category tend to get higher POR, i.e., 10.80, 22.81, 46.81, 81.82, and 109.64 for the age category of 35-44, 45-54, 55-64, 65-74, and ≥75 years old, respectively (Table 3).

## Discussion

The present study reported a national scope, population-based cross-sectional study that involved 3,911 DM subjects in Indonesia. Of them, 685 experienced ISH, indicated that the prevalence of ISH among DM subjects in this population study was 17.5%. The prevalence of ISH among DM subjects in Indonesia based on this study population was lower than the prevalence of ISH in Ghana, i.e., 37.4% based on the out-patient diabetes clinic in the teaching hospital of Tamale [14]. Similarly, as a hospital-based study, a study in Jimma, Ethiopia, found that the prevalence of ISH among DM patients was 27.6% [15]. A population-based study in district Chiem Hoa, Vietnam, observed the general elderly population aged >60 years old found a prevalence of 22.9 % [20]. Another national population-based study in the USA revealed that the prevalence of ISH in the general population was 9.4% [21]. A similar result as the current study reported by a hospital-based cohort study in Italy that observed ISH among type 2 DM and found a prevalence of 20.3 % [22].

The present study also added evidence that DM subjects with older age, i.e., ≥75 years old, was the most influential risk factor of ISH. This finding is in accordance with the previous cohort study in Italy which concluded that the mean age of type 2 DM subjects experienced ISH was 74.3 years old [22]. On the other hand, a study in Ethiopia reported that DM subjects aged ≥60 years old were the protective factor for ISH, while the age category of 47-55 years old was the risk factor with the highest OR, i.e., 2.63 [15]. Similarly, the study in Ghana showed the most frequent ISH in the DM subjects aged 50-69 years old [14]. Regarding the study population, a study in Italy and Ethiopia comparing ISH to non-ISH, including other forms of hypertension, while a study in Ghana comparing ISH to normal subjects [14,15,22]. The previous review concluded that ISH affects 10-20% of the elderly, systolic blood pressure increase with age, while diastolic blood pressure rises until the age of 50 years and then decreases after that [23]. Increase in blood pressure with age is mostly associated with arterial stiffness. Degenerative processes such as calcification and alteration of arteriosclerotic structure play a pivotal role in the formation of large artery stiffness as well as in the small vessels. Small vessel stiffness leads to the condition of peripheral vascular resistance that influences the increase of both systolic and diastolic blood pressure. The existence of large artery stiffness increases systolic blood pressure and, conversely, decreases diastolic blood pressure. The acceleration of large artery stiffness after 50 years old lead to the steeper increase of systolic blood pressure that caused the ISH condition [24].

Lipid profile leads to the process of endothelial dysfunction that affects blood pressure. HDL cholesterol tends to have inversely associated with hypertension, while non-HDL cholesterol has a positive association [25]. The present study found that HDL was inversely associated with the ISH, while in the bivariate analysis, total cholesterol showed a positive association with ISH. High HDL level, i.e., ≥40 mg/dL, was concluded as the protective factor for ISH in this study. This finding was in accordance with the Physician Health Study that reported the highest quartile of HDL level, i.e., >53 mg/dL had the lowest adjusted-RR (0.68) compared to the other quartile [26]. A study in China also reported that HDL level was inversely related to the blood pressure as well as brachial-ankle pulse-wave velocity, a marker of arterial stiffness development [25]. The atherosclerotic formation structure of the vessels also influenced by the oxidative activity of LDL cholesterol that is also inhibited by HDL [27,28]. However, a previous study in Japan reported a positive correlation between HDL and hypertension in apparently healthy people [29]. Another study revealed that a positive association between HDL and hypertension occurred in the subjects with high-level circulation CD34-positive cells, a bone marrow-derived endothelial progenitor. The level of circulating CD-34 increases as a response of the endothelial damage, therefore masking the role of HDL as endothelial protective in healthy subjects [30].

The current study also found that duration of DM, i.e., more than five years, was significantly associated with ISH, PR=1.73 (95% CI: 1.257-2.389). This finding adds the evidence that previously reported elsewhere that revealed diabetes duration and insulin treatment status were the independent predictor of ISH [31]. The progression and duration of diabetes increase complications. Duration of diabetes is associated with arterial stiffness, while arterial stiffness plays a pivotal role in ISH [31]. The gradation of DM duration as a dose-response relationship with hypertension was also described in the previous study [32]. These findings strengthen the hypothesis that diabetes precedes arterial stiffness that caused ISH; however, another study found that onset on diabetes and brachial-ankle pulse wave velocity occurred simultaneously after a longitudinal observation indicates conversely condition [33]. Indeed, there are roles of multifactor that contributed to the arterial stiffness as a major cause of ISH. Arterial stiffness is a result of degenerative processes in the extracellular matrix of elastic arteries caused by aging and many other risk factors [34].

The final model of Binary logistic regression in this study involved a history of hypertension; however, the p-value did not meet to be considered significant. The previous history of hypertension describes the condition of individuals who tend to have a genetic predisposition [35]. Hypertension is the form of the complex trait that involved multiple organs and pathways [35,36]. Comprehensive understanding of genomics, epigenomics, metabolomics, proteomics, and transcriptomics of blood pressure plays a pivotal role in the context of the previous history of hypertension [35]. Further study that observed the detailed genetic role should be conducted to elucidate the novel hypertension pathophysiology and dissect and characterize the disorder’s mechanism.

It is well established that obesity is associated with ISH [37–39]. Obesity affects the process of inflammation, cell adhesion, and coagulation that impact in the arterial stiffness [38,40]. Obesity is also related to the insulin and leptin resistance that contributes to sodium retention with concomitant cardiac output [39]. However, in this study, BMI did not significantly associate with ISH, although involved in the final model. It must be considered that the role of BMI measurement alone is inadequate for accurately predict the disease progression in DM subjects [41]. Other parameters such as body composition, total adipose mass, visceral adiposity–accumulation of intra-abdominal fat, and muscle mass should be analyzed to describe the current condition of DM subjects [41–43].

## Conclusions

The prevalence of ISH among Indonesian DM subjects in the present study was 17.5%. Older DM subjects, low HDL cholesterol, and duration of DM were associated with the ISH, suggesting that modify lipid profile, especially HDL cholesterol level, is a needful measure to delay ISH in older and duration DM subjects.

## Data Availability

The data used in this study are available from the corresponding author Mahalul Azam upon request through the email address. The data set was accessed from the RISKESDAS (Riset Kesehatan Dasar); a five-annual national basic health survey that conducted and supported by the National Institute of Health Research and Development (NIHRD), Ministry of Health, the Republic of Indonesia.The protocol and reports of the RISKESDAS is published on https://www.litbang.kemkes.go.id/laporan-riset-kesehatan-dasar-riskesdas//

## Acknowledgments

This study was supported by the Faculty of Sports Science, Universitas Negeri Semarang, Indonesia [Grant ID: 36.4.5/UN37/PPK.4.6/2020]. The RISKESDAS (Riset Kesehatan Dasar); a five-annual national basic health survey is conducted and supported by the National Institute of Health Research and Development (NIHRD), Ministry of Health, the Republic of Indonesia. The manuscript was prepared using a limited access data set obtained from the NIHRD and does not reflect the opinions or views of RISKESDAS and NIHRD. The authors thank the RISKESDAS investigators for granting permission to use their data set for the current study. The protocol and reports of the RISKESDAS is published on https://www.litbang.kemkes.go.id/laporan-riset-kesehatan-dasar-riskesdas//

